## Supplementary material for "Severity Predictors of COVID-19 in SARS-CoV-2 Variant, Delta and Omicron Period; Single Center Study": Table 1

| Table 1. Severity Classification Criteria and Therapeutic Strategy for critical COVID-19 | | |
| --- | --- | --- |
| ● Criteria for mild, severe or critical COVID-19 | | |
| 1) Moderate or severe: Oxygen demand or with risk factors: age, chronic renal failure, severe obesity, etc. 2) Critical: SpO_2_ <92% at 10L/min. oxygen via a reservoir mask | | |
| 3) Critical: Shortness of breath with respiratory rate of >30/min. | | |
| 4) Critical: Severe dyspnea due to COVID-19 pneumonia   ● Therapeutic strategy for moderate or severe COVID-19   1) Moderate: Neutralizing antibody 2) Severe: antiviral therapy: Remdesivir 5-10 days 3) Severe: Systemic steroid therapy: Dexamethasone 5-10 days 4) Antibiotics: depend on patients’ comorbidities for CAP | | |
| ● Therapeutic strategy for critical COVID-19 | | |
| 1)Mechanical ventilator | mode | pressure control |
| (primary setting) | PEEP | 10-15 mmH_2_O |
|  | Driving Pressure | 20-25 mmH_2_O |
|  | Respiratory Rate | 12-16/min. |
| (optional therapies) | Self-prone position |  |
|  | Pone position |  |
|  | ECMO |  |
| 2) Antiviral therapy | Remdesivir | 5 or 10 days |
| 3) Systemic steroid therapy | Dexamethasone | 10 days |
| 4) Anticoagulant therapy | UFH with therapeutic dose according to APTT (1.5-2 times as normal) | |
| 5) Protection for DVT | Intermittent air compression and elastic stocking | |
| 6) Antibiotics | for CAP or secondary bacterial or fungus infection |  |
| 7) Rehabilitation | early intervention by NS, PT and OT |  |
| 8) Nutrition | early intervention via tube feeding or TPN |  |
| 9) Supportive therapy | sedation, catecholamine support etc. via central venous catheter | |
| CAP: community associated pneumonia; PEEP: positive end-expiratory pressure; PS: pressure support; ECMO: Extracorporeal membrane Oxygenation; UFH: unfractionated heparin; APTT: activated partial thromboplastin time; NS: nurse; PT: physical therapist; OT: occupational therapist; TPN: total parenteral nutrition. | | |
