## Supplementary material for "Severity Predictors of COVID-19 in SARS-CoV-2 Variant, Delta and Omicron Period; Single Center Study": Table 2

Table.2 Patients’ Characteristics

|  |  | All Cases (n=141) |
| --- | --- | --- |
| Sex (Male, %) |  | 99 (70.2%) |
| Age (y.o., Median ±SD, range) |  | 58±16.1 (18-93) |
| Period from onset to hospitalization (days, Median ±SD, range) |  | 5±4.2 (0-22) |
| Period from onset to PCR positive (days, Median ±SD, range) |  | 1±2.3 (0-13) |
| Symptom (cases, %) |  |  |
|  | fever | 126 (89.4) |
|  | dyspnea | 79 (56.0) |
|  | cough | 54 (38.3) |
|  | fatigue | 48 (34.0) |
|  | sore throat | 10 7.1) |
|  | consciousness disorder | 1 (0.7) |
|  | headache | 4 (2.8) |
| Height (cm, median ±SD, range) |  | 165±10.1 (123-189) |
| Weight (kg, median ±SD, range) |  | 65±17.1 (34-121) |
| BMI (median ±SD, range) |  | 24±5.0 (14-46) |
| smoking habit (cases, %) |  | 77 (54.6) |
| vaccination (yes, %) |  | 55 (39.0) |
| comorbidities (cases, %) |  |  |
|  | respiratory disease | 20 (14.2) |
|  | cardiovascular disease | 29 (20.6) |
|  | renal disease | 40 (28.4) |
|  | continuous hemodialysis | 30 (21.3) |
|  | diabetes | 38 (27.0) |
|  | hypertension | 61 (43.3) |
|  | hyperlipidemia | 25 (17.7) |
|  | collagen diseases | 5 (3.5) |
|  | with malignant tumor | 9 (6.4) |
|  | pregnancy | 2 (1.4) |
|  | immunosuppression drugs | 7 (5.0) |

y.o.: year-old, SD: Standard Deviation, PCR: Polymerase Chain Reaction,

BMI: Body Mass Index
