## Supplementary material for "Severity Predictors of COVID-19 in SARS-CoV-2 Variant, Delta and Omicron Period; Single Center Study": Table 3

Table.3 Clinical Data in Severity Classification

|  |  | Moderate group (n=91) | Severe group (n=50) | p value |
| --- | --- | --- | --- | --- |
| Sex (Male, %) |  | 55 (60.4) | 44 (88.0) | 0.0006 |
| Age (y.o. Median ±SD, range) |  | 59±17.5 (18-93) | 57±13.3 (30-93) | 0.31 |
| Period from onset to hospitalization (days, Median ±SD, range) |  | 3±3.6 (0-13) | 8±4.2(0-22) | <0.0001 |
| Period from onset to PCR positive (days, Median ±SD, range) |  | 1±2.3 (0-13) | 2±2.3 (0-10) | 0.06 |
| Symptom (cases, %) |  |  |  |  |
|  | fever | 79 (86.8) | 47 (94.0) | 0.19 |
|  | dyspnea | 39 (42.9) | 40 (80.0) | <0.0001 |
|  | cough | 36 (39.6) | 18 (36.0) | 0.68 |
|  | fatigue | 36 (39.6) | 12 (24.0) | 0.06 |
|  | sore throat | 10 (11.0) | 0 (0) | 0.15 |
|  | consciousness disorder | 1 (1.1) | 0 (0) | 0.46 |
|  | headache | 4 (4.4) | 0 (0) | 0.13 |
| Height (cm, median ±SD, range) |  | 162.5±10.8 (123-189) | 168.5±8.0 (148-181) | 0.005 |
| Weight (kg, median ±SD, range) |  | 59.9±16.0 (34.1-121) | 74.1±16.5 (43.5-110) | <0.0001 |
| BMI (median ±SD, range) |  | 22.8±10.8 (13.8-45.5) | 26.3±8.0 (16.6-38) | 0.0001 |
| smoking habit (cases, %) |  | 42 (46.2) | 35 (70.0) | 0.007 |
| vaccination (cases, %) |  | 47 (51.6) | 39 (78.0) | 0.002 |
| comorbidities (cases, %) |  |  |  |  |
|  | respiratory disease | 14 (15.4) | 6 (12.0) | 0.58 |
|  | cardiovascular disease | 23 (25.3) | 6 (12.0) | 0.06 |
|  | renal disease | 32 (35.2) | 8 (16.0) | 0.016 |
|  | continuous hemodialysis | 24 (26.4) | 6 (12.0) | 0.046 |
|  | diabetes | 26 (28.6) | 12 (24.0) | 0.56 |
|  | hypertension | 38 (41.8) | 23 (46.0) | 0.63 |
|  | hyperlipidemia | 12 (13.2) | 13 (26.0) | 0.06 |
|  | collagen diseases | 4 (4.4) | 1 (2.0) | 0.46 |
|  | with malignant tumor | 8 (8.8) | 1 (2.0) | 0.11 |
|  | pregnancy | 1 (1.1) | 1 (2.0) | 0.67 |
|  | immunosuppression drugs | 5 (.5) | 2 (4.0) | 0.7 |
| Laboratory Data |  |  |  |  |
|  | IL-6 (pg/ml) | 52.3±121.7 (0.2-646) | 31.3±75.3 (2.1-455) | 0.16 |
|  | WBC (10^3^/μl) | 6.1±3.3 (1.5-18.7) | 6.3±6.4 (2.7-45.5) | 0.87 |
|  | CRP (mg/dl) | 5.7±6.8 (0.2-34.5) | 6.4±7.5 (0.1-37.8) | 0.2 |
|  | D-dimer (μg/ml) | 1.2±13.2 (0.5-122) | 1.1±9.6 (0.5-67.3) | 0.85 |
|  | AST (IU/l) | 34±40.2 (5-256) | 50.5±46.9 (21-252) | <0.0001 |
|  | ALT (IU/l) | 21±30.0 (3-165) | 50.5±41.3 (3-162) | <0.0001 |
|  | LDH (IU/l) | 300±136.7143-832) | 499.5±192.1 (224-1130) | <0.0001 |
|  | BUN (mg/dl) | 18±20.5 (1-91) | 19.5±19.2 (6-80) | 0.5 |
|  | Cre (mg/dl) | 1.1±3.6 (0.4-12.7) | 0.89±3.9 (0.4-14.3) | 0.06 |
|  | T.bil (mg/dl) | 0.5±0.3 (0.2-1.7 | 0.6±0.3 (0.3-1.8) | 0.16 |
| Period of whole hospitalization (days, Median ±SD, range) |  | 9±5.4 (2-36) | 16±25.6 (2-159) | <0.0001 |
| Outcome (Death, cases, %) |  | 1 (1.1) | 4 (8.0) | 0.03 |

y.o: year-old, SD: Standard Deviation, BMI: Body Mass Index, IL-6: Interleukin-6, WBC: White Blood Cell, CRP: C-reactive Protein, AST: Aspartate Aminotransferase, ALT: Alanine Aminotransferase, LDH: Lactate Dehydrogenase, Cre: Creatinine, T.bil: Total Bilirubin
