## Supplementary material for "Severity Predictors of COVID-19 in SARS-CoV-2 Variant, Delta and Omicron Period; Single Center Study": Table 4

Table 4. Therapeutic Strategy between moderate group and severe group

|  |  | Moderate Group (n=91) | Severe Group (n=50) | p value |
| --- | --- | --- | --- | --- |
| oxygen supply (cases, %) |  | 64 (70.3) | 50 (100) | <0.0001 |
| Period of oxygenation (days, Median ±SD, range) | | 4.0±6.8 (0-43) | 15±26.2 (4-164) | <0.0001 |
| mechanical ventilation (cases, %) |  | 0 (0) | 30 (60) | <0.0001 |
| Period of ventilation (days, Median ±SD, range) | | 0(0) | 7.5±24.6 (3-133) | <0.0001 |
| Tracheostomy (cases, %) |  | 0 (0) | 7 (14) | 0.0002 |
| ECMO (cases, %) |  | 0 (0) | 5 (10) | 0.0021 |
| NHF (cases, %) |  | 3 (2.1) | 23 (46) | <0.0001 |
| Period of NHF (days, Median ±SD, range) | | 9±3.5 (5-12) | 6±2.3 (1-10) | 0.11 |
| prone position (cases, %) |  | 3 (2.1) | 21 (42) | <0.0001 |
| Period of prone position (days, Median ±SD, range) | | 4±1.5 (2-5) | 4±2.8 (2-12) | 0.68 |
| Therapy | |  |  |  |
|  | steroid | 69 (48.9) | 50 (100) | 0.0002 |
|  | remdesivir | 69 (48.9) | 50 (100) | 0.0002 |
|  | tocilizumab | 0 (0) | 3 (6) | 0.02 |
|  | baricitinib | 3 (2.1) | 4 (8) | 0.22 |
|  | casirivimab | 2 (1.4) | 0 | 0.29 |
|  | molnupiravir | 16 (11.3) | 0 | 0.0016 |
|  | heparin | 61 (43.) | 50 (100) | <0.0001 |

SD: Standard Deviation, ECMO: Extracorporeal Membrane Oxygenation, NHF: Nasal High Flow
