## Supplementary material for "Severity Predictors of COVID-19 in SARS-CoV-2 Variant, Delta and Omicron Period; Single Center Study": Table 5

Table 5. Multivariable Logistic Regression

|  | OR | 95% CI | *p*-value |
| --- | --- | --- | --- |
| The period between from onset to hospitalization | 1.16 | (1.02-1.32) | 0.026 |
| BMI | 1.10 | (1.01-1.21) | 0.039 |
| Smoking habit | 3.70 | (1.41-9.68) | 0.008 |
| LDH | 1.01 | (1.00-1.01) | < 0.001 |
| OR: odds ratio; CI: confidence intervals, BMI: Body Mass Index, LDH: Lactate Dehydrogenase | |  |  |
