## Supplemental table for "Severity Predictors of COVID-19 in SARS-CoV-2 Variant, Delta and Omicron Period; Single Center Study"

Supplemental Tabel.1 Patients’ Characteristics of Different Periods (Delta Period, Omicron Period)

|  |  | Delta Period (n=89) | Omicron Period (n=52) | p value |
| --- | --- | --- | --- | --- |
| Sex (Male, %) |  | 63 (63.6%) | 36 (69.2%) | 0.85 |
| Age (y.o., Median ±SD, range) |  | 56±14.1 (18-93) | 72±15.0 (23-90) | <0.001 |
| Period from onset to admission (days, Median ±SD, range) |  | 7±4.1 (0-22) | 2±3.3 (0-13) | <0.0001 |
| Period from onset to PCR positive (days, Median ±SD, range) |  | 2±1.9 (0-10) | 1±2.9 (0-13) | 0.33 |
| Symptom (cases, %) |  |  |  |  |
|  | fever | 82 (92.1) | 44 (84.6) | 0.16 |
|  | dyspnea | 56 (62.9) | 23 (44.2) | 0.03 |
|  | cough | 35 (39.3) | 19 (36.5) | 0.74 |
|  | fatigue | 27 (30.3) | 21 (40.4) | 0.22 |
|  | sore throat | 4 (4.5) | 6 (11.5) | 0.12 |
|  | consciousness disorder | 0 (0) | 1 (1.9) | 0.19 |
|  | headache | 3 (3.4) | 1 (1.9) | 0.62 |
| height (cm, median ±SD, range) |  | 167±9.4 (142-184) | 164.6±11.1 (123-189) | 0.26 |
| Weight (kg, median ±SD, range) |  | 67.4±17.7 (37.1-121) | 60.3±14.9 (34.1-107.8) | 0.027 |
| BMI (median ±SD, range) |  | 24.7±5.1 (15.7-45.5) | 23.1±4.5 (13.8-35.7) | 0.024 |
| smoking habit (cases, %) |  | 46 (51.7) | 31 (59.6) | 0.36 |
| vaccination (yes, %) |  | 6 (6.7) | 49 (94.2) | <0.0001 |
| comorbidities (cases, %) |  |  |  |  |
|  | respiratory disease | 9 (10.1) | 11 (21.2) | 0.07 |
|  | cardiovascular disease | 7 (7.9) | 22 (42.3) | <0.0001 |
|  | renal disease | 16 (18.0) | 24 (46.2) | 0.0003 |
|  | continuous hemodialysis | 13 (14.6) | 17 (32.7) | 0.01 |
|  | diabetes | 20 (22.5) | 18 (34.6) | 0.12 |
|  | hypertension | 30 (33.7) | 31 (59.6) | 0.0027 |
|  | hyperlipidemia | 13 (14.6) | 12 (23.1) | 0.20 |
|  | collagen diseases | 2 (2.2) | 3 (5.8) | 0.28 |
|  | with malignant tumor | 5 (5.6) | 4 (7.7) | 0.63 |
|  | pregnancy | 2 (2.2) | 0 (0) | 0.28 |
|  | immunosuppression drugs | 2 (2.2) | 5 (9.6) | 0.052 |

y.o.: year-old, SD: Standard Deviation, PCR: Polymerase Chain Reaction, BMI: Body Mass Index

Supplemental Table 2. Patients’ Clinical Features by Severity Classification of Different Periods (Delta Period & Omicron Period)

|  |  | Delta Period (n=89) | | | Omicron Period (n=52) | | |
| --- | --- | --- | --- | --- | --- | --- | --- |
|  |  | Moderate (n=50) | Severe (n=39) | p value | Moderate (n=41) | Severe (n=11) | p value |
| Gender (Male, %) | | 30 (60) | 33 (84.6) | 0.011 | 25 (61.0) | 11 (100) | 0.013 |
| Age (y.o., Median ±SD, range) | | 55.5±16.4 (18-93) | 56±11.8 (30-93) | 0.95 | 72±16.2 (23-90) | 72±10.6 (44-79) | 0.76 |
| Period from onset to hospitalization (days, Median ±SD, range) | | 5±4.0 (0-13) | 8±4.1 (1-22) | 0.0003 | 1±3.1 (0-13) | 4±3.5 (0-11) | 0.025 |
| Period from onset to PCR positive (days, Median ±SD, range) | | 2±1.8 (0-8) | 2±2.2 (0-10) | 0.69 | 0±2.9 (0-13) | 2±2.9 (0-8) | 0.2 |
| Symptom (cases, %) | |  |  |  |  |  |  |
|  | fever | 45 (90) | 37 (94.9) | 0.4 | 34 (82.9) | 10 (90.9) | 0.51 |
|  | dyspnea | 25 (50) | 31 (79.5) | 0.0043 | 14 (34.1) | 9 (81.8) | 0.0047 |
|  | cough | 21 (42) | 14 (35.9) | 0.056 | 15 (36.6) | 4 (36.4) | 0.99 |
|  | fatigue | 17 (34) | 10 (25.6) | 0.39 | 19 (46.3) | 2 (18.2) | 0.09 |
|  | sore throat | 4 (8) | 0 (0) | 0.07 | 6 (14.6) | 0 (0) | 0.18 |
|  | consciousness disorder | 0 (0) | 0 (0) | N.S. | 1 (2.4) | 0 (0) | 0.6 |
|  | headache | 3 86) | 0 (0) | 0.12 | 1 (2.4) | 0 (0) | 0.6 |
| Height (cm, median, range) | | 162.5±9.6 (142-184) | 170±8.7 (148-181) | 0.029 | 162.5±12.1 (123-189) | 166±5.1 (159-176) | 0.07 |
| Weight (kg, median, range) | | 60.5±16.1 (37.1-121) | 78.7±17.3 (43.5-110) | 0.0002 | 59.4±15.8 (34.1-107.8) | 70.4±11.0 (47-85.8) | 0.12 |
| BMI (median, range) | | 22.8±5.1 (15.7-45.5) | 27.5±4.7 (16.6-38) | 0.0004 | 22.8±4.5 (13.8-35.7) | 23.9±4.5 (19-32.7) | 0.45 |
| smoking habit (cases, %) | | 20 (40) | 26 (66.7) | 0.013 | 22 (53.7) | 9 (81.8) | 0.091 |
| vaccination (yes, %) | | 5 (10) | 1 (2.6) | 0.17 | 39 (95.1) | 10 (90.9) |  |
| comorbidities (cases, %) | |  |  |  |  |  |  |
|  | respiratory disease | 6 (12) | 3 (7.7) | 0.50 | 8 (19.5) | 3 (27.3) | 0.58 |
|  | cardiovascular disease | 5 (10) | 2 (5.1) | 0.40 | 18 (43.9) | 4 (36.4) | 0.65 |
|  | renal disease | 11 (22) | 5 (12.8) | 0.26 | 21 (51.2) | 3 (27.3) | 0.16 |
|  | continuous hemodialysis | 10 (20) | 3 (7.7) | 0.10 | 14 (34.1) | 3 (27.3) | 0.67 |
|  | diabetes | 13 (26) | 7 (17.9) | 0.37 | 13 (31.7) | 5 (45.5) | 0.39 |
|  | hypertension | 14 (28) | 16 (41.0) | 0.20 | 24 (58.5) | 7 (63.6) | 0.76 |
|  | hyperlipidemia | 4 (8) | 9 (23.1) | 0.05 | 8 (19.5) | 4 (36.4) | 0.24 |
|  | collagen diseases | 2 (4) | 0 (0) | 0.21 | 2 (4.9) | 1 (9.1) | 0.59 |
|  | with malignant tumor | 4 (8) | 1 (2.6) | 0.27 | 4 (9.8) | 0 (0) | 0.28 |
|  | pregnancy | 1 (2) | 1 (2.6) | 0.86 | 0 (0) | 0 (0) | N.S. |
|  | immunosuppression drugs | 1 (2) | 1 (2.6) | 0.86 | 4 (9.8) | 1 (9.1) | 0.95 |
| oxygen supply (cases, %) | | 39 (78) | 39 (100) | 0.0018 | 25 (61.0) | 11 (100) | 0.013 |
| Period of oxygenation (days, Median ±SD, range) | | 5±8.2 (0-43) | 14.5±28.0 (4-164) | <0.0001 | 3±4.2 (0-15) | 18±20.2 (10-80) | <0.0001 |
| mechanical ventilation (cases, %) | | 0 (0) | 39 (100) | <0.0001 | 0 (0) | 5 (45.5) | <0.0001 |
| Period of ventilation (days, Median ±SD , range) | | 0 | 8±24.8 (0-133) | <0.0001 | 0 | 0±25.0 (3-79) | <0.0001 |
| Tracheostomy (cases, %) | | 0 (0) | 5 (12.8) | 0.0083 | 0 (0) | 2 (18.2) | 0.0054 |
| ECMO (cases, %) | | 0 (0) | 4(10.3) | 0.021 | 0 (0) | 1 (9.1) | 0.051 |
| NHF (cases, %) | | 2 (4) | 18 (46.2) | <0.0001 | 1 (2.4) | 5 (45.5) | <0.0001 |
| Period of NHF (days, Median ±SD, range) | | 10.5±2.1 (9-12) | 6±2.3 (1-10) | 0.034 | 5 | 6±1 (0-7) | 1 |
| prone position (cases, %) | | 2 (4) | 19 (48.7) | <0.0001 | 1 (2.4) | 2 (18.2) | 0.047 |
| Period of prone position (days, Median ±SD, range) | | 3.5±2.1 (2-5) | 4±2.9 (2-12) | 0.63 | 4 | 3 | 1 |
| Therapy | |  |  |  |  |  |  |
|  | steroid | 44 (88) | 39 (100) | 0.025 | 25 (61.0) | 11 (100) | 0.013 |
|  | remdesivir | 44 (88) | 39 (100) | 0.025 | 25 (61.0) | 11 (100) | 0.013 |
|  | tocilizumab | 0 (0) | 3 (7.7) | 0.046 | 0 (0) | 0 (0) | N.S. |
|  | baricitinib | 3 (6) | 4 (10.3) | 0.46 | 0 (0) | 0 (0) | N.S. |
|  | casirivimab | 2 (4) | 0 (0) | 0.21 | 0 (0) | 0 (0) | N.S. |
|  | molnupiravir | 0 (0) | 0 (0) | N.S. | 16 (39.0) | 0 (0) | 0.013 |
|  | heparin | 41 (82) | 39 (100) | 0.0052 | 20 (48.8) | 11 (100) | 0.0021 |
| Laboratory Data | |  |  |  |  |  |  |
|  | IL-6 | 50.6±102.5 (0.2-517) | 24.8±82.3 (2.1-455) | 0.35 | 53.5±166.6 (2.1-646) | 36±43.5 (2.1-130) | 0.25 |
|  | WBC | 5.6±2.8 (1.5-17.2) | 6.3±3.2 (2.7-14.3) | 0.094 | 7.2±3.7 (2.5-18.7) | 6.3±12.2 (2.7-45.5) | 0.85 |
|  | CRP | 6.8±7.5 (0.16-34.5) | 6.6±8.2 (0.17-37.8) | 0.83 | 3.6±5.3 (0.16-27.4) | 5.7±3.5 (0.13-10.6) | 0.39 |
|  | D-dimer | 0.96±17.4 (0.5-122) | 1.07±2.6 (0.5-12.1) | 0.35 | 1.57±2.7 (0.5-11.5) | 1.08±19.8 (0.5-67.3) | 0.88 |
|  | AST | 37±40 (5-256) | 53±41.3 (24-208) | 0.0005 | 29±40.5 (9-199) | 46±65.4 (21-252) | 0.0436 |
|  | ALT | 22±26.7 (5-117) | 56±43 (3-162) | 0.0001 | 18±33.9 (3-165) | 44±31.6 (9-116) | 0.095 |
|  | LDH | 348±26.7 (143-832) | 555±26.7 (312-1130) | <0.0001 | 270±123.7 (148-643) | 368±204.7 (224-921) | 0.023 |
|  | BUN | 29±21.1 (1-91) | 25±19.2 (6-80) | 0.29 | 21±19.4 (8-77) | 27±17.4 (15-67) | 0.25 |
|  | Cre | 0.89±3.57 (0.35-12.73) | 0.76±3.73 (0.41-13.42) | 0.2955 | 1.54±3.5 (0.4-12.7) | 1.06±4.6 (0.6-14.3) | 0.82 |
|  | T.bil | 0.5±0.4 (0.2-1.7) | 0.6±0.3 (0.3-1.3) | 0.51 | 0.5±0.3 (0.2-1.7) | 0.6±0.46 (0.4-1.8) | 0.14 |
| Period of whole hospitalization (days, Median ±SD, range) | | 9±5.4 (3-36) | 14±28.6 (2-159) | 0.0001 | 10±5.5 (2-30) | 22±10.3 (3-40) | 0.0004 |
| Death |  | 1 (2) | 1 (2.6) | 0.84 | 0 (0) | 3 (27.3) | 0.0006 |

y.o.: year-old, SD: Standard Deviation, PCR: Polymerase Chain Reaction, BMI: Body Mass Index, ECMO: Extracorporeal Membrane Oxygenation, NHF: Nasal High Flow, IL-6: Interleukin-6, WBC: White Blood Cell, CRP: C-reactive Protein, AST: Aspartate Aminotransferase, ALT: Alanine Aminotransferase, LDH: Lactate Dehydrogenase, Cre: Creatinine, T.bil: Total Bilirubin
